# Reward circuit function and treatment outcome following vALIC deep brain stimulation in treatment-resistant depression

**DOI:** 10.1101/2023.12.07.23299640

**Authors:** N. Runia, L.A. van de Mortel, C. L. C. Smith, I.O. Bergfeld, B.P. de Kwaasteniet, J. Luigjes, J. van Laarhoven, P. Notten, G. Beute, P. van den Munckhof, P.R. Schuurman, D.A.J.P. Denys, G.A. van Wingen

**Affiliations:** Amsterdam UMC location University of Amsterdam, Department of Psychiatry, Meibergdreef 9, Amsterdam, The Netherlands; Amsterdam Neuroscience, Amsterdam, The Netherlands; Amsterdam UMC location Vrije Universiteit Amsterdam, Department of Radiology and Nuclear Medicine, Boelelaan 1117, Amsterdam, The Netherlands; Cancer Center Amsterdam, Imaging and Biomarkers, Amsterdam, The Netherlands; Martini Hospital, Department of Radiology and Nuclear Medicine, Groningen, the Netherlands; Department of Psychiatry, ETZ, location Elisabeth, Tilburg, The Netherlands; Department of Neurosurgery, ETZ, location Elisabeth, Tilburg, The Netherlands; Amsterdam UMC location University of Amsterdam, Department of Neurosurgery, Amsterdam, the Netherlands

**Author notes:** These authors contributed equally to this manuscript (N. Runia, L.A. van de Mortel). Corresponding authors at: Amsterdam UMC location University of Amsterdam, Department of Psychiatry, Meibergdreef 9, 1105 AZ, Amsterdam, the Netherlands.

## Abstract

Depression is associated with abnormal functioning of the reward circuit. Several deep brain stimulation (DBS) targets for treatment-resistant depression (TRD) directly modulate white matter bundles of the reward circuit. Here we investigated whether baseline reward processing in the brain is associated with ventral anterior limb of the internal capsule (vALIC) DBS outcome and whether vALIC DBS changes neural activity in the reward circuit.

We studied fifteen patients with TRD who performed a monetary reward task during functional magnetic resonance imaging (fMRI) before vALIC DBS surgery, after DBS parameter optimization, and during a sham-controlled crossover phase. DBS devices were switched off during scanning for MRI safety reasons. Additionally, fifteen matched healthy controls were investigated twice to account for test-retest effects. We investigated brain responses to reward anticipation, loss anticipation, reward feedback and loss feedback.

Results showed that lower baseline nucleus accumbens activation during loss anticipation and higher baseline caudate nucleus and midcingulate cortex activation during reward feedback processing were associated with worse DBS outcome. No significant changes in reward processing were observed following vALIC DBS in comparison to healthy controls or after active compared to sham stimulation. Instead, increased middle frontal gyrus responses following DBS to loss feedback was associated with better DBS outcome.

These results suggest that DBS efficacy in TRD is related to individual differences in reward circuit functioning at baseline and to changes in middle frontal gyrus responses following DBS.

## 1. Introduction

Treatment-resistant depression (TRD) is commonly defined as a major depressive disorder (MDD) for which two or more prior treatments have failed to achieve clinical response (1). Patients with TRD who do not sufficiently respond to any of the conventional treatment options for MDD, may benefit from deep brain stimulation (DBS). While still in an experimental phase, DBS for severe TRD has proved to be a successful treatment, with on average 40% of patients achieving clinical response within one year (2). Common DBS targets for the treatment of TRD include the subcallosal cingulate cortex, ventral anterior limb of the internal capsule (vALIC), and medial forebrain bundle (MFB), of which the latter two are involved in the reward circuit (3–5). Dysfunction of the reward circuit and aberrations in its associated fiber bundles have previously been indicated as a possible cause for the pathological anhedonic states in depressive disorders (6–8). While not much is known about the therapeutic mechanism of vALIC DBS, prolonged stimulation of the vALIC could induce biological changes to its surrounding limbic regions within reward- and affective circuits (9–12).

Within these circuits, the nucleus accumbens (NAc) is one of the regions that can either directly be stimulated with DBS through the volume of activated tissue (13) or indirectly through connections with the vALIC (14), and could be contributing to the therapeutic changes induced by vALIC DBS. Previous studies have shown that the NAc is a key region in the brain reward circuit (15) and shows abnormal functioning in MDD (16–18). By probing the activity of the NAc during reward anticipation, we have previously shown that NAc reward processing normalizes following vALIC DBS in obsessive-compulsive disorder (9). We therefore hypothesize that such normalization of NAc activity during reward anticipation following vALIC DBS would occur in TRD as well. However, whether such normalization in reward anticipation or other measures of reward function occurs in the NAc, and to what extent this also affects other regions related to reward processing has not been investigated previously. Additionally, it is unknown whether baseline reward circuit functioning holds predictive information about future DBS treatment outcome.

In this longitudinal functional magnetic imaging (fMRI) study, we investigated reward processing in TRD patients receiving vALIC DBS treatment. Patients underwent fMRI scanning while performing a monetary reward task known to activate the NAc (19–21) before DBS implantation and following surgery and DBS parameter optimization. Matched healthy controls were included and performed the fMRI task at two time points to control for test-retest effects. To assess short-term effects of DBS de-activation after parameter optimization on reward processing, TRD patients also performed the fMRI task after double-blind periods of active and sham stimulation. We investigated the predictive properties of baseline NAc and whole-brain reward processing for DBS treatment response, and assessed longitudinal and active stimulation effects of DBS on reward processing. Additionally, we investigated whether NAc and whole-brain longitudinal changes in reward processing were related to clinical outcome.

## 2. Methods

### 2.1 Patients

We included patients with TRD as well as healthy controls in a longitudinal study followed by a randomized crossover phase (for patients only). This imaging study was an add-on to a previously reported clinical trial for vALIC DBS in TRD (13) (trial registration number: [NTR2118](https://onderzoekmetmensen.nl/nl/trial/23670)). The study was approved by the medical ethics boards of the two participating hospitals: Academic Medical Center, Amsterdam [AMC] and St Elisabeth Hospital, Tilburg [SEH]. All included patients provided written informed consent.

At inclusion, patients had to be aged 18-65 years and have a primary diagnosis of MDD according to the DSM-IV (assessed with a semi-structured clinical interview for DSM-IV disorders), an illness duration of more than 2 years, a 17-item Hamilton Depression Rating Scale (HAM-D-17) score of 18 or higher, and a Global Assessment of Function sore of 45 or lower. Additionally, patients had to be treatment-resistant, defined as a failure of at least two different classes of second-generation antidepressants, one trial of a tricyclic antidepressant, one trial of a tricyclic antidepressant with lithium augmentation, one trial of a monoamine oxidase inhibitor, and six or more sessions of bilateral electroconvulsive therapy. Patients who fulfilled the above criteria and remained stable with maintenance electroconvulsive therapy, but relapsed after discontinuation of that therapy, were also eligible. Patients had to be able to understand the consequences of the procedure (IQ >80), and capable of making choices without coercion. Exclusion criteria were a diagnosis of Parkinson’s disease, dementia, epilepsy, bipolar disorder, schizophrenia or history of psychosis unrelated to MDD, antisocial personality disorder, current tic disorder, an organic cause of depression, substance abuse during the past 6 months, unstable physical condition, pregnancy, or general contraindications for surgery.

Healthy controls were matched by age, sex, and education level. Their lifetime history of psychiatric illness and that of their first-degree relatives were negative.

### 2.2 Treatment

Four-contact leads were implanted bilaterally and connected to a neurostimulator (lead: 3389; stimulator: Activa PC/RC, Medtronic, Minneapolis, MN, USA). The electrodes were implanted with the most ventral contact at the core of the NAc and the three more dorsal contacts in the vALIC. Following a three-week recovery period, DBS setting optimization was started. The aim of DBS optimization was to maximize antidepressant effects while minimizing side effects. Standardized optimization included changing active contact points and voltage (ranging from 2.5 to 6.0 V). Pulse width and frequency were kept stable (90 microseconds and 130 or 180 Hz, respectively).

Standardized optimization ended after a response (defined as a HAM-D-17 score reduction >=50%) was reached for at least four weeks or after a maximum of 52 weeks. Details on the surgery and vALIC DBS treatment (including parameter optimization) have been described earlier (13). Individual DBS parameter settings after the standardized parameter optimization are provided in Supplementary Table 1.

### 2.3 Study Design

MRI scanning was performed three weeks prior to DBS surgery (baseline), and after DBS parameter optimization (follow-up). The aim was to keep concurrent medication stable during the optimization phase; however, psychiatrists were allowed to make changes for clinical indications (for an overview of psychotropic medications used over time see Supplementary Table 2). Healthy controls were scanned at baseline and after five months. After follow-up, the DBS group entered the double-blind randomized cross-over phase, which consisted of two blocks of one to six weeks during which DBS stimulation was on (active) or off (sham). Patients could be prematurely crossed over to the next phase while blinding was maintained if this was requested by the patient, or if the treating psychiatrist or research team deemed it clinically indicated and the HAM-D-17 score was at least 15. Concurrent medication and DBS settings were kept stable during the crossover phase. Patients again received MRI scans after both active and sham stimulation.

The severity of depressive symptoms was measured at each assessment (baseline, follow-up, active, sham) with the HAM-D-17, with higher scores indicating more severe depressive symptoms. A description of the analyses of the HAM-D-17 scores can be found in Supplementary Methods 1.2.

### 2.4 fMRI Acquisition and Monetary Reward Task

Structural and functional MRI data were collected with a 1.5T Siemens Magnetom Avanto syngo MR scanner with a transmit/receive (Tx/Rx CP) Head Coil. Conform the manufacturer’s safety instructions, the DBS devices were switched off during scanning. The devices were turned off just before entering the scanner room and turned back on immediately after leaving the scanner room. For details on the acquisition parameters, see Supplementary Methods 1.3. To probe NAc activation during fMRI data acquisition, patients performed a monetary reward task with an event-related design, which has been shown to consistently activate the NAc (19–21). For a detailed description and visualization of the task, see Supplementary Methods 1.1 and Supplementary Figure 1. In short, the task consisted of 108 trials during which one of three different cues was presented. A reward cue (blue circle) predicted a monetary reward (€0.50/€1.00/€2.00), a neutral cue (brown triangle) predicted no reward/loss, and a loss cue (pink square) predicted a monetary loss (-€0.50/-€1.00/-€2.00). Following a cue, a response window followed where a target (orange exclamation mark) was presented and patients had to respond as quickly as possible with a button press. Successful button presses within the response window led to a win or loss avoidance of the predicted monetary value depending on the presented cue, while a failure to respond within the response window led to no win, or loss of the predicted monetary value depending on the cue. The neutral cue never let to a monetary reward or loss. Subsequently, feedback was shown on screen displaying the amount of money won or lost during the trial as well as the cumulative earnings throughout the task. Reaction times were recorded for each trial. A description of the analysis of the reaction times can be found in Supplementary Methods 1.2.

### 2.4 fMRI Data Analysis

#### 2.4.1 Preprocessing

Before preprocessing a quality assessment was carried out by manually reviewing MRIQC (22) Image Quality Metrics and visual reports. (f)MRI data were preprocessed using the Statistical Parametric Mapping 12 (SPM12, https://www.fil.ion.ucl.ac.uk/) toolbox in MATLAB R2022a (The Math Works, Inc., 2022). Functional images were realigned to the mean functional image and subsequently slice-time corrected. The realignment parameters were inspected and scans with more than 4 mm (one slice thickness) movement in any direction were excluded from the analyses. Next, the T1-weighted image was coregistered to the mean functional image. The T1-weighted image was then segmented and subsequently the functional images and structural image were normalized into standardized Montreal Neurological Institute (MNI) space and resampled to 3 mm isotropic resolution. To improve the signal-to-noise ratio, the functional images were smoothed using an 8-mm full width at half maximum Gaussian kernel.

#### 2.4.2 NAc and Whole-Brain Activity

Neural activity during the monetary reward task was estimated with an event-related first-level analysis for each patient at each time-point (baseline, follow-up, active and sham stimulation) using a general linear model. Each type of cue (reward/neutral/loss x monetary value of €0.50/€1.00/€2.00), each type of feedback (reward, neutral, loss), and the six realignment parameters were modeled as separate regressors. Contrast images were created for reward anticipation (reward cues - neutral cues), loss anticipation (loss cues - neutral cues), reward feedback (reward feedback - neutral feedback), and loss feedback (loss feedback - neutral feedback) and were taken to the second-level group analyses. To assess NAc activation, one region-of-interest (ROI) mask for the left and right NAc was created based on the Automated Anatomical Labelling atlas 3 (AAL3) (23). Since the electrodes partially overlap with the NAc in the follow-up data, we created a bilateral electrode mask by first normalizing the voxel intensity of each follow-up scan using the global intensity mean, subsequently thresholding the data to only include voxels with electrode drop-out by selecting an intensity of <0.6, binarizing the data and averaging over time, and then multiplying each individual binary electrode mask to create a bilateral electrode mask averaged over the whole patient sample, which was subsequently multiplied with the bilateral NAc mask. The remaining voxel coverage of the bilateral NAc after removing the electrode artifacts is presented in Supplementary Figure 2. For the longitudinal analyses, we used this mask on both baseline and follow-up data to only include significant differences in NAc regions unaffected by the electrodes.

#### 2.4.3 fMRI Statistical Analysis

Baseline group-level analyses were performed using SPM12. To assess baseline group differences, we performed a two-sample t-test between DBS patients and healthy controls with age and sex as added covariates for each of the four contrasts on both whole-brain and ROI (NAc) level. To assess whether baseline NAc and whole-brain activity during reward processing is associated with DBS outcome at follow-up, we performed four separate linear regression analyses with the percentage change in HAM-D-17 scores (follow-up - baseline) as the outcome variable and NAc/whole-brain activity during four different contrasts (during reward anticipation, loss anticipation, reward feedback, or loss feedback) as the predictive variable, and age and sex added as covariates. Since the longitudinal change in HAM-D-17 scores violated normality assumptions, we transformed the values by taking the square root of the proportional change in HAM-D-17 scores with an added value of 1 to ensure positive values. For the resulting scores, a value of 1 represents no change, scores <1 represent decreases in HAM-D-17 scores, and scores >1 represent increases in HAM-D-17 scores following DBS.

We used voxel-wise inference for ROI analyses in the left and right NAc and corrected for multiple comparisons using a family-wise error (FWE) rate small volume correction (α<0.05). Whole-brain activation during reward processing was assessed using cluster-wise inference with a cluster-forming threshold of 0.001 and corrected for multiple comparisons using FWE correction (α<0.05).

Longitudinal group-level (patients vs. healthy controls and active vs. sham) fMRI analyses were performed using the Sandwich Estimator toolbox for SPM12 (SwE v2.2.0; http://www.nisox.org/Software/SwE) (24) (degrees of freedom type II, small sample adjustment for Wild Bootstrap resampling type C2, 999 bootstraps, unrestricted U-SwE). To assess DBS induced changes in NAc activity from baseline compared to follow-up we performed four separate mixed ANOVAs for each contrast with group (DBS vs. healthy controls) as the between-subjects factor and session (baseline vs. follow-up) as the within-subjects factor. Since the time from baseline to follow-up varied greatly within patients (mean=420.78 days, SD=159.15) and between patients and healthy controls (mean=148.00 days, SD=13.74), days since baseline was added as a time-varying covariate to the models. To assess NAc activity following active compared to sham stimulation, we carried out a repeated measures ANOVA with session (active vs. sham) as the within-subjects factor. Due to premature cross-overs, the time between sessions varied between patients (mean=20.63 days, SD=14.26). Therefore, days since baseline as well as the randomization order were added as covariates to the model.

For the longitudinal analyses (baseline vs. follow-up and active vs. sham), we used the SwE non-parametric Wild Bootstrap procedure for the ROI analysis with voxel-wise inference and statistical tests across the left and right NAc were corrected for multiple comparisons using a false discovery rate (FDR) small volume correction (α<0.05). Whole brain analyses were performed using the same SwE non-parametric Wild Bootstrap procedure with cluster-wise inference (cluster-forming threshold=0.01). Whole-brain statistical tests were corrected for multiple comparison using FWE correction (α<0.05).

To assess whether changes in NAc and whole-brain activity were related to clinical outcome, we performed separate linear regression analyses in SPM12 with the patient group only for each of the four contrasts. For these analyses we modeled the interaction between session (baseline vs. follow-up and active vs. sham) and clinical outcome (transformed percentage change in HAM-D-17 scores) while correcting for days since baseline. Results were assessed using the same statistical inference as for the baseline SPM12 analyses described above.

## 3. Results

In total, twenty-five patients and twenty-two healthy controls were included in the clinical trial, but we were unable to acquire data for each participant at all time points. For a detailed overview of the missing data see Supplementary Table 3. Apart from the missing data, we additionally excluded two patients from our baseline analysis due to unrepresentative and missing HAM-D-17 scores at follow-up, two patients due to insufficient brain coverage during fMRI acquisition, one patient due to missing onset times for the monetary reward task, one healthy control due to excessive head motion (>4 mm), and one healthy control due to a missing HAM-D-17 score at baseline, leaving 15 patients and 18 healthy controls in the baseline analyses. For the follow-up analyses, nine patients and 15 healthy controls remained after excluding one patient due to insufficient brain coverage during fMRI, one patient due to missing onset times for the monetary reward task, and two healthy controls due to excessive head motion. We included 11 patients in the active vs. sham analyses after excluding one patient due to excessive head motion.

### 3.1 Clinical Results

Demographic and clinical characteristics are presented in Table 1. Patients and healthy controls showed no statistically significant difference in sex, age and estimated IQ. At baseline, patients had significantly higher scores than healthy controls on measures of depressive symptom severity in the HAM-D-17, MADRS, and IDS-SR (all p<0.001). There was a significant reduction in HAM-D-17 score (n=9) from baseline (mean=23.70, 95%CI=18.82-28.51) to follow-up (mean=12.80, 95%CI=7.28-18.28; t=-2.375, p=0.033), and patients had a significantly lower HAM-D-17 score (n=11) after active stimulation (mean=14.70, 95%CI=10.08-19.37) compared to sham stimulation (mean=22.00, 95%CI=18.41-25.59; t=-3.380, p=0.0079).

**Table 1.** Descriptive characteristics of DBS patients and healthy controls.

| Characteristics | Baseline |  | p-value<sup>†</sup> | Baseline – follow-up |  | p-value<sup>†</sup> | Active-sham |
| --- | --- | --- | --- | --- | --- | --- | --- |
|  | DBS | HC |  | DBS | HC |  | DBS |
| Sample size | 15 | 18 |  | 9 | 15 |  | 11 |
| Sex, No. (%) |  |  |  |  |  |  |  |
| Female | 12 (80%) | 11 (61%) | n.s. | 8 (87.5%) | 9 (60%) | n.s. | 8 (73%) |
| Male | 3 (20%) | 7 (39%) |  | 1 (12.5%) | 6 (40%) |  | 3 (27%) |
| Age at inclusion |  |  |  |  |  |  |  |
| mean (SD) | 52.9 (7.9) | 53.2 (8.0) | n.s. | 51.6 (8.4) | 52.9 (8.3) | n.s. | 53.3 (4.9) |
| median (IQR)** | 55.0 (11.0) | 55.0 (7.0) |  | 50.0 (6.0) | 56.0 (6.0) |  |  |
| Estimated IQ, mean (SD) | 98.0 (15.6) | 102.0 (13.5) | n.s. | 96.6 (17.4) | 104.0 (14.0) | n.s. | 96.4 (13.2) |
| Baseline HAM-D-17 score |  |  |  |  |  |  |  |
| mean (SD) | 23.1 (5.6) | 0.9 (1.3) | * | 23.7 (6.3) | 0.7 (1.3) | * | 22.4 (6.19) |
| median (IQR) | 23 (8.5) | 0.0 (1.0) |  | 23.0 (12.0) | 0.0 (1.0) |  |  |
| Baseline MADRS score |  |  |  |  |  |  |  |
| mean (SD) | 34.0 (6.3) | 0.8 (1.3) | * | 33.9 (6.8) | 0.9 (1.4) | * | 33.8 (5.8) |
| median (IQR) | 36.0 (7) | 0.0 (1.8) |  | 36.0 (7.0) | 0.0 (2.0) |  |  |
| Baseline IDS-SR score |  |  |  |  |  |  |  |
| mean (SD) | 49.5 (9.6) | 3.2 (2.7) | * | 47.9 (9.9) | 3.0 (2.3) | * | 46.7 (11.5) |
| median (IQR) | 51.0 (14.0) | 3.0 (4.0) |  | 47.0 (11.0) | 3.0 (4.0) |  |  |
| Age at MDD onset, mean (SD) |  |  |  |  |  |  |  |
| Self-report | 28.7 (16.0) | - |  | 29.9 (17.0) | - |  | 30.5 (15.3) |
| Diagnosis | 37.1 (10.4) | - |  | 36.8 (12.1) | - |  | 38.3 (11.8) |
| No. of past medications |  |  |  |  |  |  |  |
| mean (SD) | 10.0 (3.0) | - |  | 10.7 (3.6) | - |  | 10.5 (3.1) |
| median (IQR) |  |  |  |  |  |  | 10.0 (2.5) |
| No. of past ECT series |  |  |  |  |  |  |  |
| mean (SD) | 2.2 (1.1) | - |  | 2.0 (1.2) | - |  | 1.8 (1.1) |
| median (IQR) | 2.0 (2.0) | - |  |  | - |  | 1.0 (1.5) |
| No. of past ECT sessions |  |  |  |  |  |  |  |
| mean (SD) | 52.8 (49.2) | - |  | 56.7 (60.5) | - |  | 50.2 (55.5) |
| median (IQR) | 47 (30) | - |  | 49.0 (37.0) | - |  | 42.0 (28.5) |
| Years since MDD onset, diagnosis, mean (SD) | 15.8 (8.8) | - |  | 14.8 (8.8) | - |  | 15.0 (10.7) |
| MDD episodes, No. (%) |  |  |  |  |  |  |  |
| 1 | 6 (40%) | - |  | 4 (50%) | - |  | 6 (55%) |
| 2 | 2 (13%) | - |  | 2 (25%) | - |  | 3 (27%) |
| >2 | 7 (47%) | - |  | 2 (25%) | - |  | 2 (18%) |

Abbreviations: DBS, Deep Brain Stimulation; HC, healthy controls; SD, standard deviation; IQ, intelligence quotient; HAM-D, Hamilton Rating Scale for Depression; MADRS, Montgomery-Åsberg Depression Rating Scale; IDS-SR, Inventory of Depressive Symptomatology–Self-report; MDD, major depressive disorder; ECT, electroconvulsive therapy.
<sup>†</sup> p-values were obtained with appropriate parametric and non-parametric tests.
\* Significant differences between patients and healthy controls (p<0.001).
\*\* median indicated when normality assumptions were violated.

### 3.2 Behavioral Results

For both comparative analyses there was a significant main effect of condition for reaction time (baseline vs. follow-up: F(2,107.96)=3.942, p=0.008; sham vs. active: F(2,48)=5.018, p=0.01), with average reaction times after reward and loss cues being significantly shorter compared to average reaction times after neutral cues. Additionally, the baseline vs. follow-up analysis revealed a significant interaction effect between group and session (F(1,112.42)=7.038, p= 0.009). Post-hoc comparisons showed that there was no significant difference in average reaction times between patients and healthy controls at baseline. However, average reaction times in healthy controls significantly decreased at follow-up compared to baseline, whereas there was no significant difference in patients. There were no significant interaction effects with condition (p>0.05). There was also no significant difference in average reaction times after active compared to sham stimulation (p>0.05) and no significant interaction between those sessions and condition (p>0.05).

### 3.3 fMRI results

#### 3.3.1 Baseline

Group comparisons did not show any statistically significant differences between patients and healthy controls at baseline in NAc and whole-brain activity during reward anticipation, loss anticipation, reward feedback, or loss feedback (p_fwe-corrected_>0.05).

Regression analyses within the patient group between baseline activity and DBS outcome showed that lower NAc activity during loss anticipation was significantly associated with worse outcome (p_fwe-corrected_=0.025, see Figure 1). Additionally, we found that higher midcingulate (MNI: 18,-1,35; cluster extent: 40 voxels; p_fwe-corrected_=0.029, see Figure 2) and caudate nucleus (MNI: - 3,17,17;cluster extent: 72 voxels; p_fwe-corrected_=0.002, see Figure 3) activity in reward feedback was significantly associated with worse outcome.

**Figure 1.**
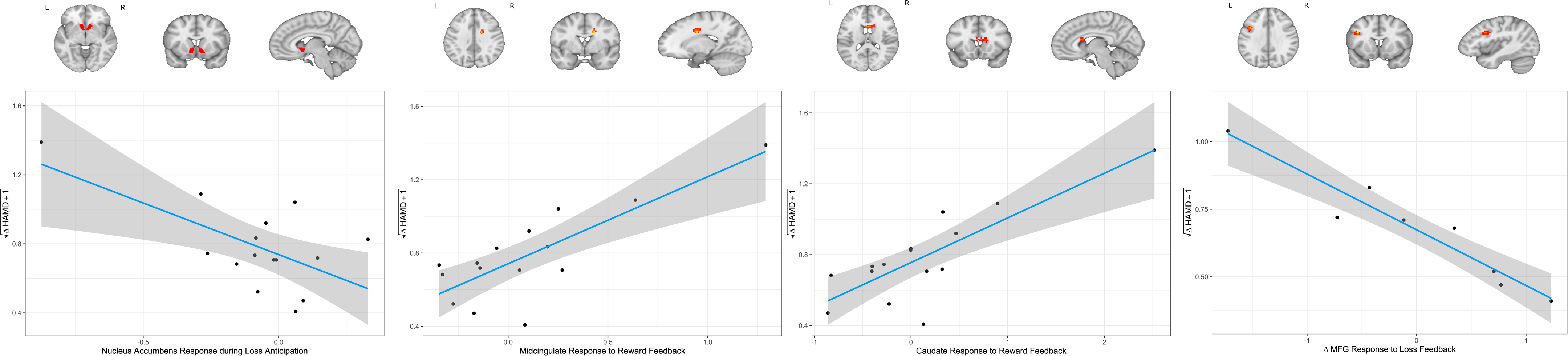
Negative association between nucleus accumbens response during loss anticipation at baseline and DBS outcome. Results from ROI-based regression analysis revealed that higher responses during loss anticipation led to a better outcome in vALIC DBS (p_FWE-corrected_=0.025). HAMD=Hamilton-Depression Rating Scale.

**Figure 2:**
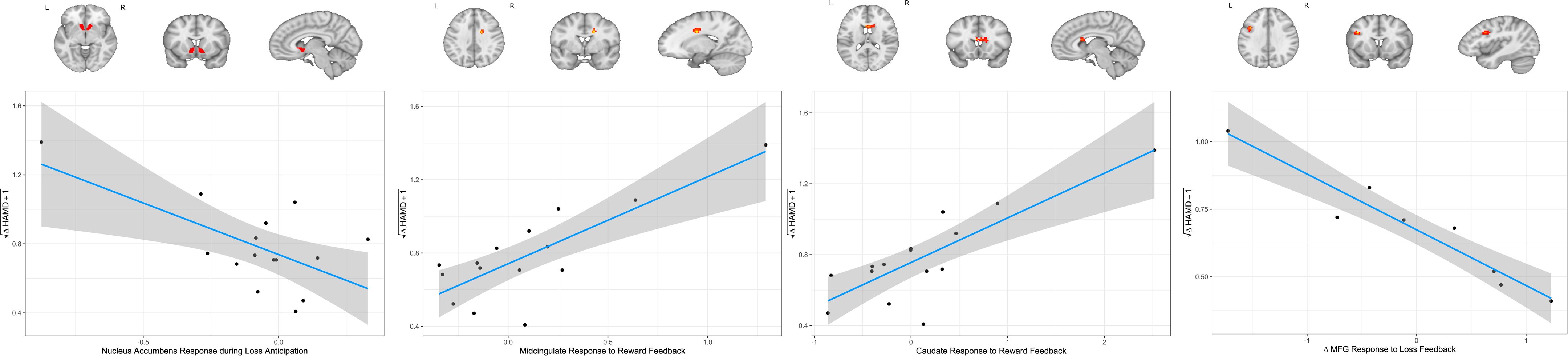
Positive association between right midcingulate response to reward feedback at baseline and DBS outcome. Whole-brain regression analysis revealed a statistically significant cluster in the right midcingulate gyrus where lower responses to reward feedback led to a better outcome in vALIC DBS (p_FWE-corrected_=0.029). HAMD=Hamilton-Depression Rating Scale.

**Figure 3:**
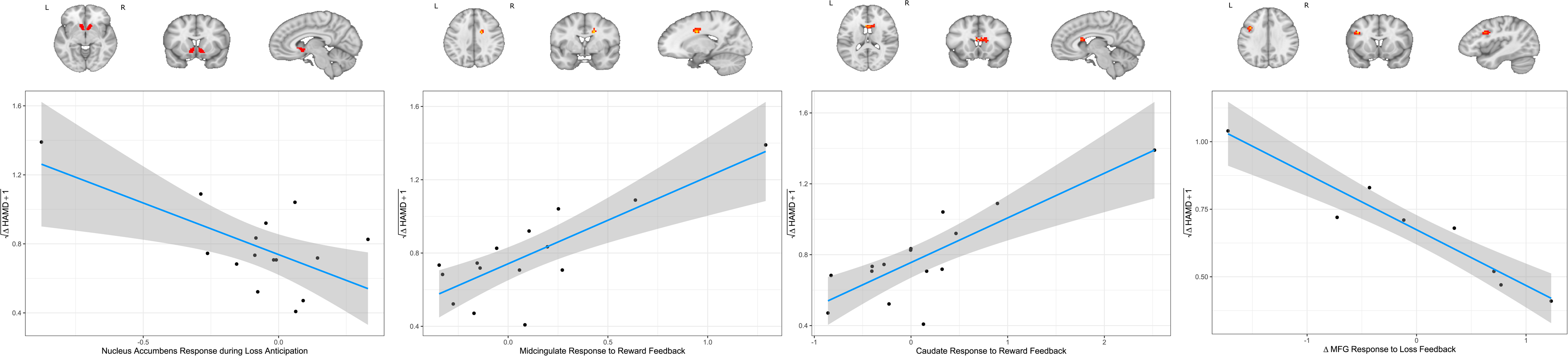
Positive association between right caudate nucleus response to reward feedback at baseline and DBS outcome. Whole-brain regression analysis revealed a statistically significant cluster in the right caudate nucleus, where lower responses to reward feedback led to a better outcome in vALIC DBS (p_FWE-corrected_=0.002). HAMD=Hamilton-Depression Rating Scale.

One of the patients had an atypical large increase in HAM-D-17 score following DBS combined with largely deviating brain responses during reward processing at baseline. The HAM-D-17 change score of this patient was statistically not deemed to be an outlier. Nonetheless, we investigated whether the associations between activity in these clusters and DBS outcome remained significant without this patient. Within the clusters, associations remained significant for both the caudate nucleus (p=0.02) and midcingulate (p=0.02), but not for the NAc (p>0.05). However, the clusters did not remain statistically significant when we reperformed the whole-brain analyses after excluding this patient.

#### 3.3.2 Baseline vs. Long-Term Follow-up

Neither the whole-brain analysis nor the NAc ROI-analysis showed significant session x group interaction effects (p_FWE/FDR-corrected_>0.05) for any of the four different contrasts. None of the contrasts showed a significant effect of session on the whole-brain level (p_FWE-corrected_>0.05) or specifically in the NAc (p_FDR-corrected_>0.05). Additionally, no significant main effects of group were found after whole-brain analysis (p_FWE-corrected_>0.05). However, ROI-analysis of the NAc showed a trend towards a significant main effect of group during reward feedback (left: p_FDR-corrected_=0.073, right: p_FDR-corrected_=0.073). An exploratory post-hoc comparison showed that patients exhibited decreased right and left NAc activity during reward feedback compared to healthy controls (left: p_FDR-corrected_=0.039, right: p_FDR-corrected_=0.039). The other contrasts did not show significant main effects of group in the NAc (p_FDR-corrected_>0.05).

One patient in the longitudinal data did not have a representative HAM-D-17 score at follow up, and we thus assessed if activity changes over time were dependent on the clinical outcome within the group of eight remaining patients. Whole-brain analysis on the activity change (follow-up – baseline) and DBS outcome revealed that an increase in left middle frontal gyrus (MFG) activity during loss feedback was significantly associated with better clinical outcome (MNI: −39,11,32; cluster extent: 32 voxels; p_FWE-corrected_=0.037, see Figure 4). NAc ROI-analysis for loss feedback did not show an association between activity change and clinical outcome (p_FWE-corrected_>0.05), and none of the other contrasts showed any significant associations between activity change in the NAc or on the whole-brain level and clinical outcome (p_FWE-corrected_>0.05).

**Figure 4:**
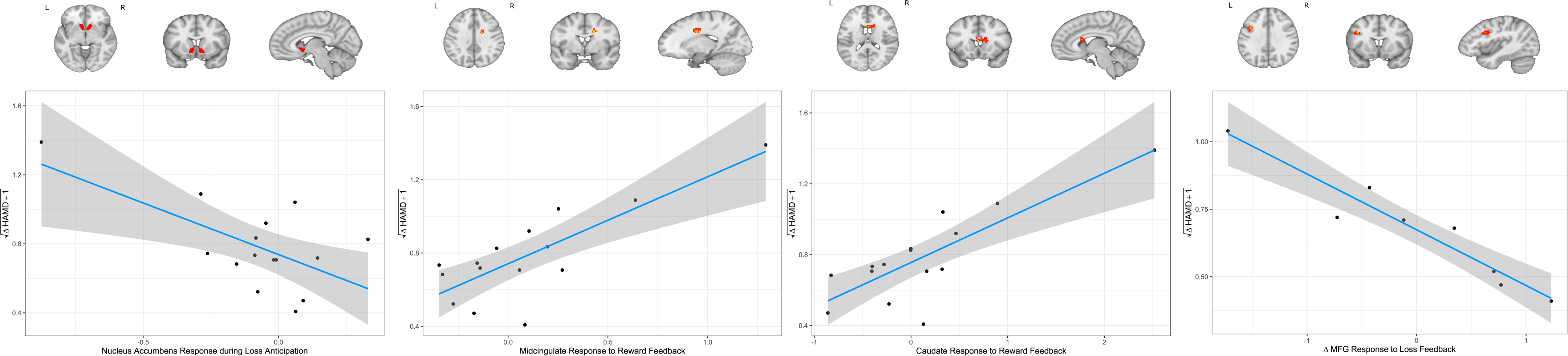
Association between longitudinal changes in the left middle frontal gyrus response to loss feedback and DBS outcome. Whole-brain regression analysis revealed an association between changes in the left MFG response to loss feedback and HAM-D-17 change following vALIC DBS treatment, with an increase in left MFG response to loss feedback being associated with higher clinical improvement (p_FWE-corrected_=0.037). HAMD=Hamilton Depression Rating Scale, MFG=middle frontal gyrus.

#### 3.3.3 Active vs. Sham DBS

Whole-brain analysis and NAc ROI-analysis of the cross-over phase (active vs. sham) showed no significant effects of session (p_FWE/FDR-corrected_>0.05) for any of the different contrasts. Additionally, there were no significant associations between activity change in the NAc or on the whole-brain level and clinical outcome for any of the four contrasts (p_FWE_>0.05).

## 4. Discussion

This study aimed to assess the effect of vALIC DBS on reward processing in the NAc and on the whole-brain level, and to study both baseline and longitudinal associations between brain activity during reward processing and DBS outcome. The results showed that higher caudate nucleus and midcingulate cortex activation during reward feedback processing and lower NAc activation during loss anticipation were associated with worse DBS outcome. Although DBS resulted in a significant decrease in depressive symptoms, we found no significant effect of DBS on changes in reward processing when the whole group of patients was compared to healthy controls, nor after a period of active compared to sham stimulation. Though within the patient group, we observed a significant association between increased MFG response to loss feedback following DBS and a better DBS outcome in patients.

The brain regions found to be associated with DBS outcome (the NAc, midcingulate cortex, and the caudate nucleus) have previously shown to be involved in reward processes and reward-based decision making (25–27). In addition, these regions have previously been implicated in the neurobiology of TRD (28–30), and have consistently shown hypoactivity during reward feedback in MDD and relations to anhedonia (17, 31, 32). While we did not find a significant difference in brain activity in these regions between patients and controls at baseline, our findings imply that functional variation in the NAc, midcingulate cortex, and caudate nucleus within the patient group may still be relevant for the outcome of DBS.

Despite a significant reduction in depression symptoms after (active) DBS, patients did not show changes in brain reward circuit regions following long-term DBS or after active compared to sham stimulation. This result is in contrast with our previous study in obsessive-compulsive disorder, in which we reported that DBS restores aberrant NAc function during reward processing (9). This discrepancy could possibly be explained by different underlying biological disease mechanisms, but may also be due to differences in study design, as the study in obsessive-compulsive disorder compared patients following one year of active stimulation and after one week of DBS cessation. Nonetheless, since a large body of literature suggests that MDD is associated with aberrant reward processing in the brain (e.g. 8, 17, 33) and we specifically aimed to target the reward circuit by stimulation of the vALIC, it was surprising to find no DBS induced changes in reward processing. As we did not include a scale that specifically measured anhedonia, we cannot be entirely sure that vALIC DBS reduced anhedonia. Since anhedonia is specifically linked to activity in the reward circuit (e.g. 32), this could explain why we did not find reward circuit modulation after DBS. However, given the substantial reduction in depressive symptoms in our sample, it seems unlikely that DBS did not have any influence on anhedonia.

But while there were no significant changes over time in reward processing when we compared the entire group of DBS patients to healthy controls, we did find an association between MFG activity during loss feedback and DBS outcome. Specifically, our results imply that MFG activity increases following DBS lead to better DBS outcome. The MFG has previously shown involvement in reward and loss processing (34, 35), may genetically and functionally be involved in depression (17, 36), and is an important target for repetitive transcranial magnetic stimulation treatment of depression (37). While we found this association in only a small group of patients and thus requires further investigation, it corroborates previous findings on the involvement of the MFG in depression and its clinical treatments.

Overall, we observed quite remarkable variation in recorded brain responses during the various reward conditions. While this facilitated the detection of associations between neural responses and DBS outcome, we did not find overall differences between DBS patients and healthy controls at baseline. The high variation recorded in the small sample of this study may have limited our power in detecting group differences. Additionally, one of the patients in this study had a deviating HAM-D-17 change score combined with more extreme brain responses during reward processing, which was largely driving the associations between brain responses during reward processing and DBS outcome. Although these associations remained significant in the caudate nucleus and midcingulate without this patient, the association between NAc responses to loss anticipation and DBS outcome disappeared, and we were not able to detect significant clusters when removing the patient from the analyses. While this limits the robustness of our findings, it could also be that smaller associations were not detectable with the reduced power after removing this patient altogether. Nonetheless, the deviating HAM-D-17 change score of this patient was not a statistical outlier and high variation in responses to reward cues is common in regions such as the NAc in MDD, with the most deviating responses correlating with anhedonia and suicidal ideation severity (38). To the best of our knowledge, no study has previously investigated the role of brain reward circuit functioning as a predictor for DBS outcome. Thus, we cannot conclude whether the recorded responses to different reward cues in this patient was an exception and significant outlier, or whether this largely deviating response holds clinical relevance for associations with DBS outcome.

An important limitation of this study is the limited number of patients that could be included in the longitudinal analysis, which may have affected our results and their replicability in similar future studies (39). This study was part of a randomized controlled clinical trial, and the required sample size was calculated based on the required power to determine the therapeutic efficacy of vALIC DBS (13). The effects of DBS on fMRI measures were secondary outcomes of the study for which we unfortunately had less data (and thus power) to detect reward processing changes when compared to healthy controls. Due to this lack of power, this study also did not control for additional factors such as medication use. Nevertheless, the sample size is relatively large for an fMRI study with TRD patients undergoing DBS. Future studies powered with fMRI outcomes as one of the primary outcome measures, have to reveal whether DBS may have smaller effects on reward processing and to investigate how clinical factors affect the outcome of DBS and its effects on reward processing. Another limitation is that we were unable to investigate acute DBS effects on brain regions involved in reward processing as the DBS systems had to be switched off during scanning due to safety reasons. Future studies using DBS systems which can be safely turned on during scanning could determine whether acute effects of DBS on reward processing in the brain play a role in its working mechanism.

In summary, we found associations between baseline activity in the midcingulate cortex, caudate nucleus, and NAc during reward processing and DBS outcome in TRD, but no general short-term or long-term effects of DBS on reward processing compared to healthy controls. Instead, we observed a significant association between MFG response increases to loss feedback and a better DBS outcome. These findings have to be replicated in larger samples to determine whether reward processing could be relevant for our understanding and optimization of DBS treatment in patients with TRD.

## Data Availability

All data produced in the present study are available upon reasonable request to the authors

## Acknowledgments

Funding: This investigator-initiated study was funded by Medtronic Inc (25 DBS systems, in kind) and a research grant from ZonMw (nr. 171201008).

## Competing interests

The authors declare the following financial interests/personal relationships which may be considered as potential competing interests: This investigator-initiated study was funded by Medtronic Inc (25 DBS systems, in kind) and a grant from ZonMw (nr. 171201008). The funders had no role in the design, execution, and analysis of the study, nor in writing of the manuscript or the decision to publish. Nora Runia, Isidoor Bergfeld, Pepijn van den Munckhof, P. Richard Schuurman, Damiaan Denys, and Guido van Wingen currently execute an investigator-initiated clinical trial on deep brain stimulation for depression, which is funded by Boston Scientific (24 DBS systems in kind) and a grant of ZonMw (nr. 636310016). P. Richard Schuurman acts as consultant for Boston Scientific and Medtronic on educational events. All other authors do not declare any conflicts of interest.

(Supplementary information is available at MP’s website)

## Notes

### Clinical Trial

NTR2118

### Clinical Protocols

https://doi.org/10.1001/jamapsychiatry.2016.0152

### Author Declarations

Ethics committee of Amsterdam University Medical Center, location AMC gave ethical approval for this work Ethics committee of St. Elisabeth Hospital, Tilburg gave ethical approval for this work

### Summary of Updates

Significant changes include: Results section updated to clarify exlusion of participants, manuscript updated throughout to mention DBS being switched off during scanning. Individual DBS parameters and clinical response added to supplemental files.

## References

1. Gaynes BN, Lux L, Gartlehner G, Asher G, Forman-Hoffman V, Green J, et al. Defining treatment-resistant depression. Depression and anxiety. 2020;37(2):134–45.

2. Bergfeld IO, Figee M. Deep Brain Stimulation for Depression. Fundamentals and Clinics of Deep Brain Stimulation: Springer; 2020. p. 279–90.

3. Coenen VA, Schlaepfer TE, Goll P, Reinacher PC, Voderholzer U, Van Elst LT, et al. The medial forebrain bundle as a target for deep brain stimulation for obsessive-compulsive disorder. CNS spectrums. 2017;22(3):282–9.

4. Coenen VA, Schlaepfer TE, Maedler B, Panksepp J. Cross-species affective functions of the medial forebrain bundle—Implications for the treatment of affective pain and depression in humans. Neuroscience & Biobehavioral Reviews. 2011;35(9):1971–81.

5. Coenen VA, Schlaepfer TE, Sajonz B, Döbrössy M, Kaller CP, Urbach H, Reisert M. Tractographic description of major subcortical projection pathways passing the anterior limb of the internal capsule. Corticopetal organization of networks relevant for psychiatric disorders. NeuroImage: Clinical. 2020;25:102165.

6. Dillon DG, Gonenc A, Belleau E, Pizzagalli DA. Depression is associated with dimensional and categorical effects on white matter pathways. Depression and anxiety. 2018;35(5):440–7.

7. Henderson SE, Johnson AR, Vallejo AI, Katz L, Wong E, Gabbay V. A preliminary study of white matter in adolescent depression: relationships with illness severity, anhedonia, and irritability. Frontiers in psychiatry. 2013;4:152.

8. Höflich A, Michenthaler P, Kasper S, Lanzenberger R. Circuit mechanisms of reward, anhedonia, and depression. International Journal of Neuropsychopharmacology. 2019;22(2):105–18.

9. Figee M, Luigjes J, Smolders R, Valencia-Alfonso C-E, Van Wingen G, De Kwaasteniet B, et al. Deep brain stimulation restores frontostriatal network activity in obsessive-compulsive disorder. Nature neuroscience. 2013;16(4):386–7.

10. Park HR, Kim IH, Kang H, McCairn KW, Lee DS, Kim B-N, et al. Electrophysiological and imaging evidence of sustained inhibition in limbic and frontal networks following deep brain stimulation for treatment refractory obsessive compulsive disorder. PLoS One. 2019;14(7):e0219578.

11. Runia N, Bergfeld IO, de Kwaasteniet BP, Luigjes J, van Laarhoven J, Notten P, et al. Deep brain stimulation normalizes amygdala responsivity in treatment-resistant depression. Molecular psychiatry. 2023;28(6):2500–7.

12. Schlaepfer TE, Bewernick BH, Kayser S, Hurlemann R, Coenen VA. Deep brain stimulation of the human reward system for major depression—rationale, outcomes and outlook. Neuropsychopharmacology. 2014;39(6):1303–14.

13. Bergfeld IO, Mantione M, Hoogendoorn ML, Ruhé HG, Notten P, van Laarhoven J, et al. Deep brain stimulation of the ventral anterior limb of the internal capsule for treatment-resistant depression: a randomized clinical trial. JAMA psychiatry. 2016;73(5):456–64.

14. Gutman DA, Holtzheimer PE, Behrens TE, Johansen-Berg H, Mayberg HS. A tractography analysis of two deep brain stimulation white matter targets for depression. Biological psychiatry. 2009;65(4):276–82.

15. Bayassi-Jakowicka M, Lietzau G, Czuba E, Steliga A, Waśkow M, Kowiański P. Neuroplasticity and Multilevel System of Connections Determine the Integrative Role of Nucleus Accumbens in the Brain Reward System. Int J Mol Sci. 2021;22(18).

16. Liu R, Wang Y, Chen X, Zhang Z, Xiao L, Zhou Y. Anhedonia correlates with functional connectivity of the nucleus accumbens subregions in patients with major depressive disorder. NeuroImage: Clinical. 2021;30:102599.

17. Yang X, Su Y, Yang F, Song Y, Yan J, Luo Y, Zeng J. Neurofunctional mapping of reward anticipation and outcome for major depressive disorder: a voxel-based meta-analysis. Psychological medicine. 2022;52(15):3309–22.

18. Zhou B, Chen Y, Zheng R, Jiang Y, Li S, Wei Y, et al. Alterations of static and dynamic functional connectivity of the Nucleus Accumbens in patients with Major Depressive Disorder. Frontiers in Psychiatry. 2022;13:877417.

19. Knutson B, Adams CM, Fong GW, Hommer D. Anticipation of increasing monetary reward selectively recruits nucleus accumbens. Journal of Neuroscience. 2001;21(16):RC159-RC.

20. Knutson B, Fong GW, Adams CM, Varner JL, Hommer D. Dissociation of reward anticipation and outcome with event-related fMRI. Neuroreport. 2001;12(17):3683–7.

21. Knutson B, Westdorp A, Kaiser E, Hommer D. FMRI visualization of brain activity during a monetary incentive delay task. Neuroimage. 2000;12(1):20–7.

22. Esteban O, Birman D, Schaer M, Koyejo OO, Poldrack RA, Gorgolewski KJ. MRIQC: Advancing the automatic prediction of image quality in MRI from unseen sites. PloS one. 2017;12(9):e0184661.

23. Rolls ET, Huang C-C, Lin C-P, Feng J, Joliot M. Automated anatomical labelling atlas 3. Neuroimage. 2020;206:116189.

24. Guillaume B, Hua X, Thompson PM, Waldorp L, Nichols TE, Initiative AsDN. Fast and accurate modelling of longitudinal and repeated measures neuroimaging data. NeuroImage. 2014;94:287–302.

25. Balleine BW, Delgado MR, Hikosaka O. The role of the dorsal striatum in reward and decision-making. Journal of Neuroscience. 2007;27(31):8161–5.

26. Burton AC, Nakamura K, Roesch MR. From ventral-medial to dorsal-lateral striatum: neural correlates of reward-guided decision-making. Neurobiology of learning and memory. 2015;117:51–9.

27. Vogt BA. Midcingulate cortex: structure, connections, homologies, functions and diseases. Journal of chemical neuroanatomy. 2016;74:28–46.

28. Amiri S, Arbabi M, Kazemi K, Parvaresh-Rizi M, Mirbagheri MM. Characterization of brain functional connectivity in treatment-resistant depression. Progress in Neuro-Psychopharmacology and Biological Psychiatry. 2021:110346.

29. Sun J, Ma Y, Guo C, Du Z, Chen L, Wang Z, et al. Distinct patterns of functional brain network integration between treatment-resistant depression and non treatment-resistant depression: A resting-state functional magnetic resonance imaging study. Progress in Neuro-Psychopharmacology and Biological Psychiatry. 2023;120:110621.

30. Wu QZ, Li DM, Kuang WH, Zhang TJ, Lui S, Huang XQ, et al. Abnormal regional spontaneous neural activity in treatment-refractory depression revealed by resting-state fMRI. Human brain mapping. 2011;32(8):1290–9.

31. Pizzagalli DA, Holmes AJ, Dillon DG, Goetz EL, Birk JL, Bogdan R, et al. Reduced caudate and nucleus accumbens response to rewards in unmedicated individuals with major depressive disorder. American Journal of Psychiatry. 2009;166(6):702–10.

32. Wacker J, Dillon DG, Pizzagalli DA. The role of the nucleus accumbens and rostral anterior cingulate cortex in anhedonia: integration of resting EEG, fMRI, and volumetric techniques. Neuroimage. 2009;46(1):327–37.

33. Zhang W-N, Chang S-H, Guo L-Y, Zhang K-L, Wang J. The neural correlates of reward-related processing in major depressive disorder: a meta-analysis of functional magnetic resonance imaging studies. Journal of affective disorders. 2013;151(2):531–9.

34. Dugré JR, Dumais A, Bitar N, Potvin S. Loss anticipation and outcome during the Monetary Incentive Delay Task: a neuroimaging systematic review and meta-analysis. PeerJ. 2018;6:e4749.

35. Knutson B, Bhanji JP, Cooney RE, Atlas LY, Gotlib IH. Neural responses to monetary incentives in major depression. Biological psychiatry. 2008;63(7):686–92.

36. Liu X, Hou Z, Yin Y, Xie C, Zhang H, Zhang H, et al. CACNA1C gene rs11832738 polymorphism influences depression severity by modulating spontaneous activity in the right middle frontal gyrus in patients with major depressive disorder. Frontiers in Psychiatry. 2020;11:73.

37. Pommier B, Vassal F, Boutet C, Jeannin S, Peyron R, Faillenot I. Easy methods to make the neuronavigated targeting of DLPFC accurate and routinely accessible for rTMS. Neurophysiologie Clinique/Clinical Neurophysiology. 2017;47(1):35–46.

38. Misaki M, Suzuki H, Savitz J, Drevets WC, Bodurka J. Individual variations in nucleus accumbens responses associated with major depressive disorder symptoms. Scientific reports. 2016;6(1):21227.

39. Turner BO, Paul EJ, Miller MB, Barbey AK. Small sample sizes reduce the replicability of task-based fMRI studies. Communications biology. 2018;1(1):62.

