## Supplementary Information for "Reward circuit function and treatment outcome following vALIC deep brain stimulation in treatment-resistant depression"

*Content:*

1. *Supplementary Methods*
   1.1 *Monetary Reward Task*
   1.2 *Clinical and behavioral statistical analyses*
   1.3 *MRI Acquisition Parameters*

*Supplementary Table 1*

*Supplementary Table 2*

*Supplementary Table 3*

*Supplementary Figure 1*

*Supplementary Figure 2*

### *1.* Supplementary Methods

#### *1.1 Monetary Reward Task*

To probe NAc activation during fMRI data acquisition participants performed a Monetary Reward task with an event-related design, which has been shown to consistently activate the NAc (1-3). The task consisted of 108 trials which each lasted between 4750 and 8750 ms. A trial started with the presentation of one of three different cues (750 ms). A reward cue (blue circle) predicted a monetary reward, a neutral cue (brown triangle) predicted no reward/loss, and a loss cue (pink square) predicted a monetary loss. Additionally, each cue had either one, two, or three white stripes, indicating a reward or loss of €0.50, €1.00 or €2.00, respectively. The cue was followed by a delay (1000 or 3000 ms), after which a time window followed where a target was presented (orange exclamation mark). The target duration was either long (72 trials) or short (36 trials) and was determined by adding 400 ms to the shortest reaction time during a set of practice trials and subtracting 150 ms from the shortest reaction time during the practice trials, respectively. The order of the long and short target duration was randomly generated, with the same order used for each participant and each session. During the time window in which the target was presented participants had to respond as quickly as possible with a button press. Successful button presses within the response window led to a win or loss avoidance of the predicted monetary value depending on the presented cue, while a failure to respond within the response window led to no win, or loss of the predicted monetary value depending on the cue. The neutral cue never let to a monetary reward or loss. Subsequently, after a short delay, feedback was shown on screen (1000 ms) displaying the amount of money won or lost during the trial as well as the cumulative earnings throughout the task. After another delay (1000 or 3000 ms) the next trial started. This design ensured that all participants performed approximately equal. Reaction times were recorded for each trial. For a visualization of the task see Supplementary Figure 1.

#### *1.2 Clinical and behavioral statistical analyses*

Clinical and behavioral data were analyzed with R version 4.0.3 (R Core Team, 2020). HAM-D-17 scores at baseline and follow-up were compared with a linear mixed-effects model analysis with HAM-D-17 score as the dependent variable and session (baseline vs. follow-up) and days since baseline as independent variables. In addition, for the active/sham stimulation comparison we performed a linear mixed-effects model analysis with HAM-D-17 as the dependent variable and session (active vs. sham), days since baseline, and randomization order as independent variables.

Reaction times during the monetary reward task at baseline and follow-up were analyzed using a linear mixed-effects model. Reaction time was included as the dependent variable and group (DBS vs. healthy controls), session (baseline vs. follow-up), condition (reward vs. neutral vs. loss), and their interactions were included as independent variables. Additionally, days since baseline was included in the model as an independent variable. Furthermore, for the active/sham stimulation comparison we performed a linear mixed-effects model analysis with reaction time as the dependent variable, and session (active vs. sham), condition (reward vs. neutral vs. loss), and their interaction were included as independent variables. In addition, days since baseline and randomization order were included in the model as independent variables.

#### *1.3 MRI Acquisition Parameters*

Echo-planar images with transversal orientation were acquired during the monetary reward task (repetition time=2000 ms, echo-time=30 ms, flip angle=90°, matrix=64x64, number of slices=25, slice thickness=4.0 mm, slice gap=10%, slice order=interleaved ascending (odd first), field of view=230x230, voxel-size= 3.6x3.6x4.0 mm). 390 volumes were acquired with a total duration of 13 minutes. For anatomical reference, a three dimensional single shot T1-weighted image with sagittal orientation was acquired (repetition time=1900 ms, echo-time=3.08 ms, flip angle=15°, matrix =512x512, number of slices=192, slice thickness=1.0 mm, slice gap=50%, field of view=256x256, voxel-size=0.5x0.5x1.0 mm).

**Supplementary Table 1. Individual patient HAM-D-17 scores (baseline and follow-up), percentage HAM-D-17 reduction, and DBS parameter settings after standardized parameter optimization (follow-up) (Activa PC/RC, Metronic).**

| | | | | Left electrode | | | | | | | | | Right electrode | | | | | | | | |
|---|---|---|---|---|---|---|---|---|---|---|---|---|---|---|---|---|---|---|---|---|---|
| Patient (in random order) | HAM-D-17 baseline | HAM-D-17 follow-up | HAM-D-17 percentage reduction (%) | Case | Ventral contact | Ventromedial contact | Dorsolateral contact | Dorsal contact | Electric potential (V) | Pulse width (µs) | Frequency (Hz) | Electric current (mA)* | Case | Ventral contact | Ventromedial contact | Dorsolateral contact | Dorsal contact | Electric potential (V) | Pulse width (µs) | Frequency (Hz) | Electric current (mA)* |
| 1[#] | 26 | 14 | 46.2 | + | | - | | | 2.9 | 90 | 160 | 2.59 | + | | - | | | 2.9 | 90 | 160 | 2.76 |
| 2[‡] | 16 | 6 | 62.5 | + | | | - | | 4.3 | 90 | 180 | 4.86 | + | | | - | | 4.3 | 90 | 180 | 5.30 |
| 3[#] | 16 | 8 | 50.0 | + | - | - | - | - | 4.0 | 90 | 130 | 7.91 | + | - | - | - | - | 4.0 | 90 | 130 | 7.75 |
| 4[#†‡] | 18 | 4 | 77.8 | + | | - | - | | 5.5 | 90 | 180 | -- | + | | - | - | | 5.5 | 90 | 180 | -- |
| 5[#†‡] | 23 | 16 | 30.4 | + | | - | | | 5.2 | 60 | 130 | 5.32 | + | | - | | | 5.2 | 60 | 130 | 4.19 |
| 6[#‡] | 18 | 10 | 44.4 | + | | - | - | | 4.2 | 60 | 130 | 6.05 | + | | - | - | | 4.2 | 60 | 130 | 5.71 |
| 7[#] | 26 | 22 | 15.4 | + | - | - | - | | 4.0 | 150 | 140 | 9.02 | + | | - | - | - | 4.0 | 150 | 140 | 8.68 |
| 8[#†‡] | 30 | 14 | 53.3 | + | | - | - | | 3.5 | 90 | 180 | 5.82 | + | | - | - | - | 6.0 | 90 | 180 | 11.44 |
| 9[#] | 15 | 29 | -93.3 | + | - | - | | | 3.6 | 90 | 180 | 5.14 | + | - | - | | | 3.6 | 90 | 180 | 4.91 |
| 10[#†‡] | 31 | 16 | 48.4 | + | - | - | | | 4.5 | 90 | 130 | 7.29 | + | - | - | | | 4.5 | 90 | 130 | 7.82 |
| 11[†‡] | 16 | 17 | -6.3 | + | | - | - | | 5.4 | 90 | 180 | 9.43 | + | | - | - | | 5.4 | 90 | 180 | -- |
| 12[#] | 22 | 15 | 31.8 | + | | | - | - | 5.2 | 90 | 130 | 7.45 | + | | | - | - | 3.5 | 90 | 130 | -- |
| 13[#†‡] | 33 | 9 | 72.7 | + | | - | - | - | 7.3 | 90 | 180 | 14.20 | + | | - | - | - | 7.3 | 90 | 180 | 14.09 |
| 14[#†‡] | 20 | 10 | 50.0 | + | | | - | - | 4.3 | 90 | 180 | 5.63 | + | | | - | - | 4.3 | 90 | 180 | 6.16 |
| 15[‡] | 22 | 16 | 27.3 | + | | | - | - | 2.5 | 60 | 130 | 5.42 | + | | | - | - | 2.5 | 60 | 130 | 3.94 |
| 16[#†] | 24 | 26 | -8.3 | + | | | - | - | 6.7 | 90 | 180 | 10.79 | + | | | - | - | 6.7 | 90 | 180 | 10.93 |
| 17[‡] | 19 | 26 | -36.8 | + | | - | - | - | 3.8 | 90 | 190 | 6.05 | + | | - | - | - | 3.8 | 90 | 190 | 5.99 |
| 18[#†] | 18 | 3 | 83.3 | + | | - | - | | 5.4 | 90 | 180 | 7.64 | + | | - | - | | 5.4 | 90 | 180 | -- |
| 19[#] | 27 | 22 | 18.5 | + | | - | - | | 4.0 | 60 | 130 | 6.01 | + | | - | - | | 4.0 | 60 | 130 | 5.85 |

*The electric current was a non-adjustable parameter, resulting from the electric potential settings and the electrical resistance of the circuit. If a value is missing, resistance measures were unavailable.

[#] Included in the baseline analyses

[†] Included in the baseline vs. follow-up analyses

[‡] Included in the active vs. sham analyses

**Supplementary Table 2. Number of patients using psychotropic medication over time**

| | | Baseline – follow-up (n=9) | | Active-sham (n=11) | |
|---|---|---|---|---|---|
| | | Baseline | Follow-up | Baseline | Follow-up |
| **Antidepressant** | Combination | 1 | 2 | 1 | 3 |
| | Single | 5 | 1 | 6 | 2 |
| | None | 3 | 6 | 4 | 6 |
| **Benzodiazepine** | Single | 3 | 3 | 5 | 5 |
| | None | 6 | 6 | 6 | 6 |
| **Antipsychotic** | Single | 5 | 5 | 6 | 5 |
| | None | 4 | 4 | 5 | 6 |
| **Lithium** | Single | 0 | 0 | 1 | 1 |
| | None | 9 | 9 | 10 | 10 |
| **Anxiolytic** | Single | 0 | 1 | 0 | 1 |
| | None | 9 | 8 | 11 | 10 |
| **Anti-epileptic** | Single | 0 | 0 | 1 | 1 |
| | None | 9 | 9 | 10 | 10 |
| **Antihistaminic** | Single | 1 | 1 | 1 | 1 |
| | None | 8 | 8 | 10 | 10 |
| **Opioid** | Single | 0 | 0 | 1 | 1 |
| | None | 9 | 9 | 10 | 10 |
| **Sympathomimetic** | Single | 1 | 1 | 2 | 2 |
| | None | 8 | 8 | 9 | 9 |

**Supplementary Table 3. Reasons for missing fMRI data.**

| Patient | Baseline fMRI data available | Complete baseline/ follow-up fMRI data | Complete cross-over phase fMRI data | Reason for missing data |
|---|---|---|---|---|
| 1 | No | No | No | MRI coil was unavailable at baseline. Because there was no baseline data it was decided to not collect fMRI data at follow-up and during the cross-over phase. However, from this patient onwards fMRI data was collected at the following assessments despite missing baseline data. |
| 2 | No | No | Yes | MRI coil was unavailable at baseline. |
| 3 | No | No | Yes | MRI coil was unavailable at baseline. |
| 4 | Yes | Yes | Yes | |
| 5 | Yes | Yes | No | Patient was deemed unfit to participate in the cross-over phase due to unstable clinical status. |
| 6 | Yes | No | No | Drop-out due to non-response. |
| 7 | No | No | No | Patient was treated with MRI-incompatible vagus nerve stimulation. |
| 8 | Yes | No | No | Drop-out due to non-response. |
| 9 | Yes | Yes | Yes | |
| 10 | Yes | No | No | Drop-out due to non-response. |
| 11 | Yes | Yes | Yes | |
| 12 | Yes | No | No | Follow-up time deviated too much from protocol (2.5 years). Patient was deemed unfit to participate in the cross-over phase due to unstable clinical status. |
| 13 | Yes | Yes | Yes | |
| 14 | Yes | No | No | Patient withdrew from participation after the baseline assessment due to somatic complaints. |
| 15 | Yes | Yes | Yes | |
| 16 | Yes | No | No | Drop-out due to non-response. |
| 17 | Yes | Yes | Yes | |
| 18 | Yes | Yes | No | Unknown |
| 19 | No | No | Yes | Baseline fMRI data was not collected due to back complaints at the time. |
| 20 | Yes | Yes | No | fMRI data collection was terminated due to an anxiety attack during one of the cross-over assessments. |
| 21 | Yes | Yes | Yes | |
| 22 | Yes | No | No | At follow-up the buttons necessary for the fMRI task did not work. At one of the assessments during the cross-over phase the reward task did not work. |
| 23 | Yes | No | Yes | fMRI data collection was terminated due to an anxiety attack at the follow-up assessment |
| 24 | Yes | Yes | Yes | |
| 25 | Yes | No | No | Task did not work during the follow-up assessment. Patient was deemed unfit to participate in the cross-over phase due to unstable clinical status. |
| **Healthy control** | | **Complete baseline/ follow-up fMRI data** | | **Reason for missing data** |
| 1 | Yes | Yes | | |
| 2 | Yes | Yes | | |
| 3 | Yes | Yes | | |
| 4 | Yes | Yes | | |
| 5 | Yes | Yes | | |
| 6 | Yes | Yes | | |
| 7 | Yes | Yes | | |
| 8 | Yes | No | | Participant withdrew from participation after the baseline assessment |
| 9 | Yes | Yes | | |
| 10 | Yes | Yes | | |
| 11 | Yes | Yes | | |
| 12 | Yes | Yes | | |
| 13 | Yes | No | | Unknown |
| 14 | Yes | Yes | | |
| 15 | Yes | No | | Unknown |
| 16 | No | No | | MRI-incompatible elbow pin |
| 17 | No | No | | fMRI data collection was terminated due to an anxiety attack |
| 18 | Yes | Yes | | |
| 19 | Yes | Yes | | |
| 20 | Yes | Yes | | |
| 21 | Yes | Yes | | |
| 22 | Yes | Yes | | |

*Abbreviations:* (f)MRI, (functional) magnetic resonance imaging.

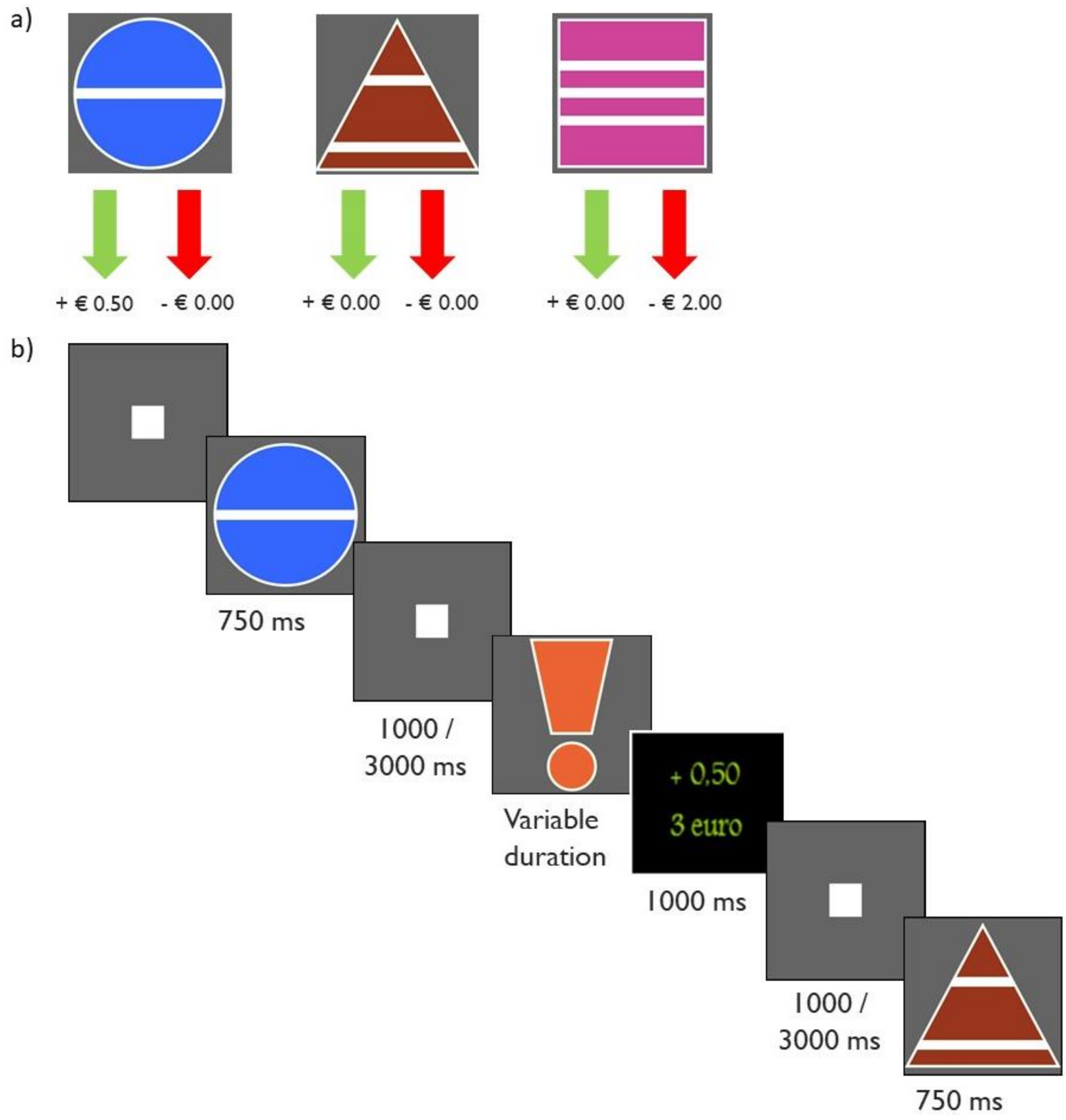


**Supplementary Figure 1. Monetary Reward Task.** a) The three different cues involved in the task: a reward cue (blue circle) predicting monetary reward, a neutral cue (brown triangle) predicting no reward/loss, and a loss cue (pink square) predicting monetary loss. Each cue had either one, two, or three white stripes, indicating a reward or loss of €0.50, €1.00 or €2.00, respectively. The green and red arrows show the result of hit and miss trial respectively for each cue. b) Task design. The trial started with the presentation of a cue followed by a target (orange exclamation mark). During target presentation the participant had to press a button as quickly as possible. Lastly, feedback was shown on screen displaying the amount of money won or lost during the trial as well as the cumulative earnings throughout the task.

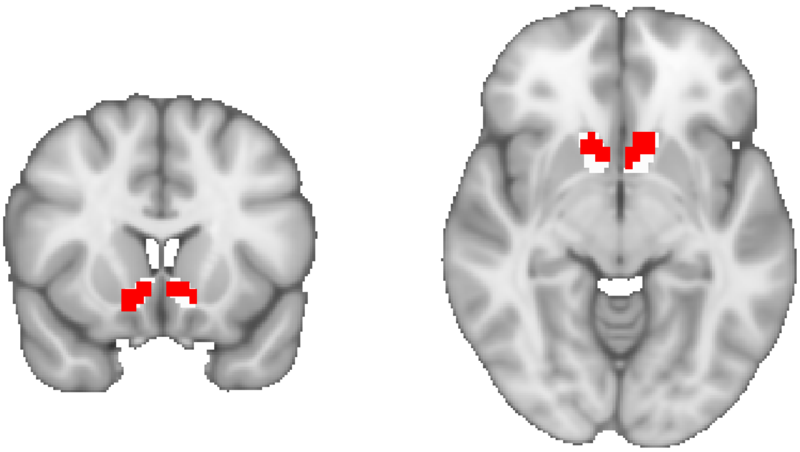

**Supplementary figure 2.** Remaining voxel coverage of the bilateral NAc (red) after removing DBS electrode artifacts as compared to the original NAc mask (white).

### References


1. Knutson B, Adams CM, Fong GW, Hommer D. Anticipation of increasing monetary reward selectively recruits nucleus accumbens. Journal of Neuroscience. 2001;21(16):RC159-RC.

2. Knutson B, Fong GW, Adams CM, Varner JL, Hommer D. Dissociation of reward anticipation and outcome with event-related fMRI. Neuroreport. 2001;12(17):3683-7.

3. Knutson B, Westdorp A, Kaiser E, Hommer D. FMRI visualization of brain activity during a monetary incentive delay task. Neuroimage. 2000;12(1):20-7.
